# Whole Genome Sequencing Reveals a *RET* Enhancer Risk Haplotype Associated with Hirschsprung Disease in Mowat Wilson Syndrome

**DOI:** 10.64898/2026.03.19.26348831

**Authors:** Sydney Collins, Ibrahim Bah, Ryan Pysar, David Mowat, Tychele N. Turner, Sumantra Chatterjee

**Author notes:** Co-corresponding authors: Send All correspondence to: Sumantra Chatterjee, Ph.D., Centre for Human Genetics & Genomics, New York University Grossman School of Medicine 435 East 30^th^ Street, Room 1012, New York, NY 10016, E and Tychele N. Turner, Ph.D., Department of Genetics, Washington University School of Medicine, 4523 Clayton Avenue, Campus Box 8232, St. Louis, MO 63110, E. Co-first authors (Sydney Collins, Ibrahim Bah).

## Abstract

Mowat Wilson syndrome (MWS) is a rare neurodevelopmental disorder caused by mostly heterozygous loss-of-function variants in *ZEB2*. Affected individuals show considerable wide variability in clinical presentation. In particular, Hirschsprung disease (HSCR) occurs in only a subset of patients, suggesting that additional genetic factors may modify disease penetrance. To investigate this possibility, we performed whole-genome sequencing of two parent-child trios in which the probands carried pathogenic *de novo ZEB2* variants but differed in enteric phenotype: one individual with MWS and long-segment HSCR and another with MWS without HSCR. In both probands, the *ZEB2* variants represent the primary causative genomic diagnosis, and no additional rare coding variants or excess copy-number burden provided a clear alternative explanation for HSCR. Phasing of a previously defined 10 single nucleotide polymorphisms(SNPs) *RET* enhancer haplotype revealed inheritance of a high-risk haplotype in the proband with HSCR, whereas the proband without HSCR carried only low-risk haplotypes on both chromosomes. To place these findings in a developmental context, we analysed single-cell transcriptomic data from the developing human fetal gut and neocortex. *ZEB2* and *RET* show overlapping expression in enteric neural crest progenitors and neuroblasts but minimal overlap in the developing neocortex, indicating that reduced *RET* dosage is likely to have tissue-specific effects in the enteric nervous system. Together, these results support a model in which common regulatory variation at *RET* modifies HSCR penetrance in the setting of *ZEB2* haploinsufficiency. More broadly, our findings illustrate how whole-genome sequencing can reveal regulatory modifiers that contribute to variable expressivity in ostensibly monogenic disorders

## INTRODUCTION

Mowat Wilson syndrome (MWS) (OMIM #235730) is a congenital Mendelian disorder characterized by a distinctive facial appearance together with variable moderate to severe intellectual disability (ID), epilepsy, Hirschsprung disease (HSCR), and multiple congenital anomalies, including genital anomalies (particularly hypospadias), congenital heart disease, agenesis of the corpus callosum, and structural malformations of the hippocampus (Mowat et al. 1998; Garavelli et al. 2017; Ivanovski et al. 2018). The prevalence of MWS is currently unknown, although estimates range from 1 in 50,000 to 1 in 100,000 live births, and it is likely that the syndrome remains under diagnosed, particularly in patients without HSCR (Zweier et al. 2002). Most reported individuals with MWS are sporadic cases, so the recurrence risk is considered low, although families with affected siblings have been described (Garavelli and Mainardi 2007). All published MWS cases to date harbour heterozygous mutations or deletions in *ZEB2*, which encodes a transcription factor of the Zfh1 family of two handed zinc finger and homeodomain proteins (Cacheux et al. 2001; Wakamatsu et al. 2001).

A key unresolved aspect of MWS is the striking phenotypic variability in body systems involved, even though all genetically confirmed cases share a similar proposed pathogenic mechanism -haploinsufficiency in *ZEB2*. This variability is seen not only between unrelated individuals but also within families that carry the same *ZEB2* variant. For example, two siblings with the p.R695X mutation showed the characteristic facial gestalt, *HSCR*, and agenesis of the corpus callosum, yet were discordant for congenital heart disease and ocular coloboma (McGaughran et al. 2005). In another family, two affected sisters with the same *ZEB2* mutation had similarly severe hypoplasia of the corpus callosum, but HSCR was present in only one sibling (McGaughran et al. 2005). Likewise, (Schuster et al. 2022) reported two siblings with the identical p.Arg343* variant, one of whom had sensorineural hearing loss and HSCR, whereas the other lacked both features. These observations strongly suggest that, beyond the primary *ZEB2* lesion, additional genetic variants modulate the expressivity and organ specific involvement of MWS.

By assessing constraint in control individuals (without MWS), it has been observed that *ZEB2* is highly intolerant to loss-of-function variation, with only 1 loss-of-function variant observed in 141,456 control individuals (Karczewski et al. 2020). *ZEB2* has also recently been shown to carry significant protein-coding *de novo* variants (DNVs) in individuals with neurodevelopmental disorders (NDDs) (Kaplanis et al. 2020). Since all MWS patients have mutations in *ZEB2*, we estimated the prevalence of MWS using data from this recent study (Kaplanis et al. 2020) where 28 out of 31,058 children with NDDs had a protein-coding disruptive DNV affecting *ZEB2* (likely individuals with MWS). Since NDDs have a prevalence of 13.87% (Boyle et al. 2011), we estimate that the prevalence of MWS is ∼1/10,000, which is higher than previous estimates.

Approximately 40 percent of individuals with MWS present with Hirschsprung disease (HSCR; congenital colonic aganglionosis; OMIM #142623) (Ivanovski et al. 2018). When present, HSCR is often considered a useful diagnostic clue for MWS and has been proposed to contribute to the male skewed sex ratio observed in MWS (2.13:1, male:female) (Mowat et al. 2003). HSCR is a congenital functional obstruction of the gut that results from developmental defects of the enteric nervous system (ENS) (Fries and Chatterjee 2024). It affects approximately 1 in 5,000 live births in individuals of European ancestry and is defined by variable cranio caudal aganglionosis of the myenteric and submucosal plexuses, with secondary megacolon and motility dysfunction. Clinically, HSCR is classified by length of aganglionosis into short segment (∼80% cases), long segment (∼15%), and total colonic aganglionosis (5%), and is characterized by high heritability (∼85%), marked genetic heterogeneity by segment length, and sex dependent reduced penetrance of Mendelian and oligogenic forms (Badner et al. 1990; Fries and Chatterjee 2024).

Given the high frequency (40%) of HSCR among individuals with MWS, and the well-established polygenic and oligogenic architecture of isolated HSCR, we hypothesized that canonical HSCR risk alleles act as genetic modifiers of the MWS phenotype. Specifically, we considered three, not mutually exclusive, possibilities for why a person with MWS might also develop HSCR: (i) the presence of additional protein coding variants in known HSCR genes (Tilghman et al. 2019) or genes expressed in the ENS (Chatterjee et al. 2023), (ii) presence of the previously defined high risk enhancer haplotype at the primary HSCR gene *RET*, which encodes a receptor tyrosine kinase essential for ENS development (Chatterjee et al. 2016; Chatterjee et al. 2021; Chatterjee et al. 2023), or (iii) other, as yet unidentified, genetic modifiers.

To test this hypothesis in a tractable setting, we performed whole-genome sequencing (WGS) on two parent child trios, one in which the proband had MWS without HSCR and one in which the proband had MWS with HSCR. Beyond the *ZEB2* mutations, we found that the proband with HSCR carries the previously defined ten SNP high risk RET enhancer haplotype, whereas the proband without HSCR does not. These data provide direct evidence that a common, incompletely penetrant HSCR risk haplotype at *RET* can modify the clinical expression of a Mendelian *ZEB2* disorder. More broadly, our findings illustrate how systematic application of WGS in apparently monogenic syndromes can uncover layered architectures of genetic risk that explain variable expressivity and tissue specific involvement, and they argue that integration of Mendelian and complex disease genetics will be essential for accurate prognosis and precision management in neurocristopathies such as MWS.

## RESULTS

### Family Pedigree

We extracted blood-derived DNA from two MWS trios, each composed of unaffected parents and an affected child. Proband 1 (MWS1.1) was diagnosed with long segment Hirschsprung disease in addition to other classical MWS features (**Figure 1A**). Proband 2 (MWS2.1) did not have Hirschsprung disease (**Figure 1A**). Targeted Sanger sequencing identified a frameshift variant (p.(Lys284Asnfs*30*)) in MWS1.1, and a stop-gained variant (p.(Arg671)) in MWS2.1.

**Figure 1.**
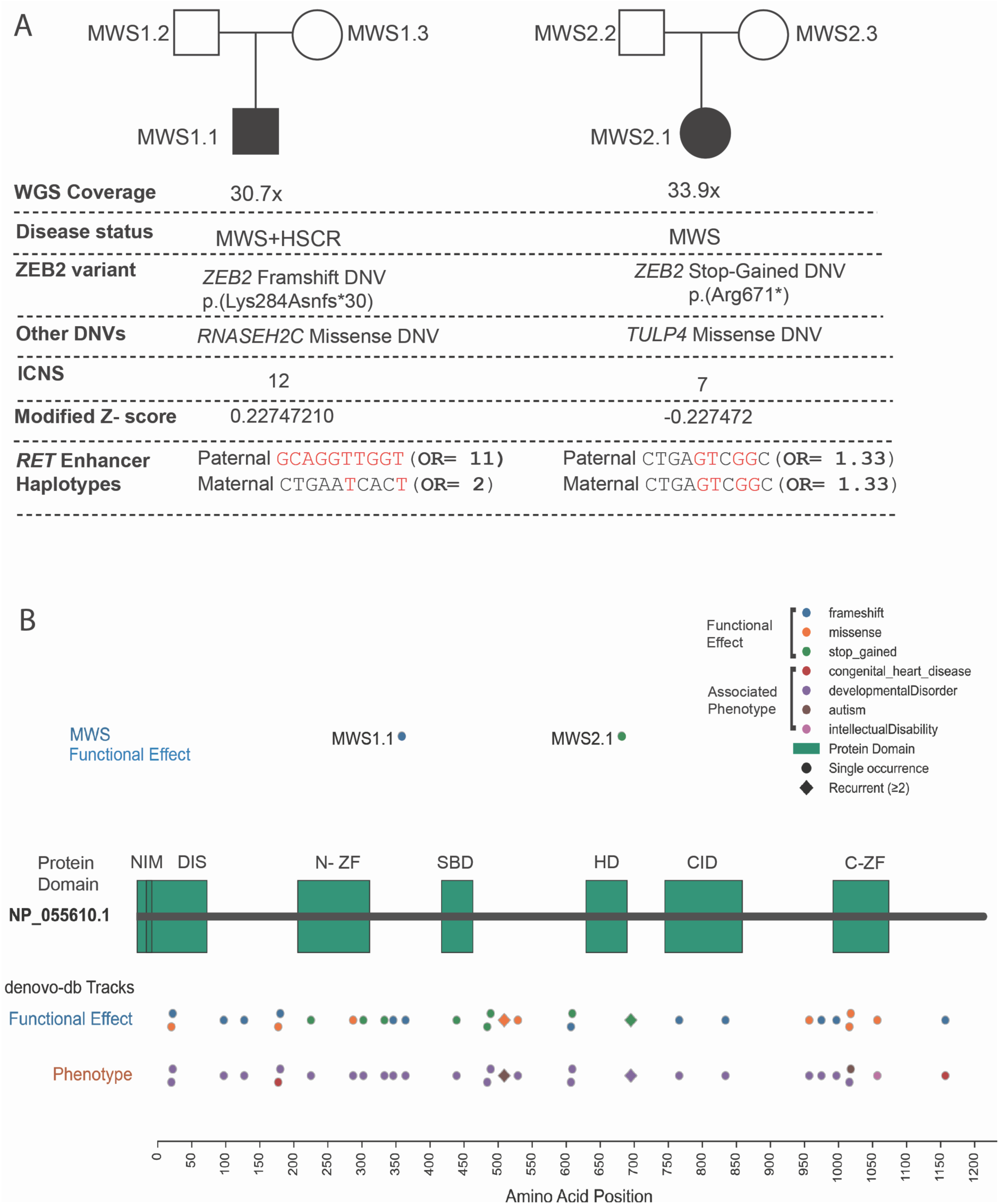
Genetic characterization of two Mowat Wilson syndrome trios and comparison with previously reported *ZEB2 de novo* variants. **(A)** Pedigrees and whole-genome sequencing results for the MWS1 and MWS2 families. Filled symbols indicate affected probands. For each proband, the *ZEB2* de novo variant (DNV), additional protein-altering DNVs, copy-number burden, and phased *RET* enhancer haplotypes are shown. MWS1.1 carries a frameshift *ZEB2* DNV (p.Lys284Asnfs30) and presents with MWS and HSCR, whereas MWS2.1 carries a stop-gained *ZEB2* DNV (p.Arg671) and presents with MWS alone. Haplotype reconstruction reveals inheritance of a high-risk paternal *RET* enhancer haplotype in MWS1.1, whereas MWS2.1 carries low-risk haplotypes from both parents. **(B)** Location of *ZEB2* variants identified in MWS1.1 and MWS2.1 relative to previously reported DNVs compiled from *denovo-db* and mapped onto the ZEB2 protein. Variants are colored by predicted functional effect (frameshift, missense, stop-gained), and associated neurodevelopmental phenotypes reported in *denovo-db* are indicated by color-coded markers. Symbol shape distinguishes single from recurrent variants. The proband variants occur within regions of the protein that harbor multiple previously reported pathogenic DNVs associated with neurodevelopmental disorders.

### ZEB2 Variant Discovery in MWS1.1 and MWS2.1

Using *Exomiser* (Smedley et al. 2015) on variant calls from our whole-genome sequencing data we identified *ZEB2* loss-of-function variants in both individuals (**Figure 1A**). In MWS1.1, the variant was a frameshift (p.(Lys284Asnfs*30)) and in MWS2.1 the variant was a stop-gained (p.(Arg671*)). These were the same variants as had been previously discovered with Sanger sequencing.

### DNVs in MWS1.1 and MWS2.1

We identified 69 DNVs in MWS1.1 and 70 in MWS2.1 (**Figure 1A, Supplementary Table S1, Supplementary Table S2**). Beyond the *ZEB2* DNVs described above, we identified two additional missense variants. In MWS1.1, the missense variant was in *RNASEH2C* (NP_115569.2:p.Arg22Pro). In MWS2.1, the missense variant was in *TULP4* (NP_064630.2:p.Pro693Leu).

To assess the potential relevance of these variants to enteric nervous system development, we examined their expression patterns in available single-cell datasets from the developing human fetal gut (Elmentaite et al. 2021). *RNASEH2C* is broadly expressed across multiple non-neuronal cell types, including smooth muscle, epithelial, and extracellular matrix-associated cell populations, but does not show enriched expression within enteric neural crest-derived lineages. Similarly, *TULP4* transcripts are not detectably expressed in ENS precursor populations in the developing fetal gut. These expression patterns make it unlikely that variants in either gene would specifically disrupt ENS development or contribute directly to HSCR.

Although we cannot completely exclude the possibility that these variants may have subtle or context-dependent biological effects, the absence of expression within ENS progenitor populations and the lack of prior association with neurocristopathies suggest that they are unlikely to represent primary drivers of the HSCR phenotype in these probands. Instead, these findings support the interpretation that the *ZEB2* pathogenic variants represent the principal Mendelian lesion in both individuals.

### Assessing Copy Number Burden in MWS1.1 and MWS2.1

To assess overall copy number burden, we utilized the ICNS feature of CNPI (Ustanik and Turner 2025) (**Supplementary Table S3**). This analysis enables discovery of overall excess of CN burden within an individual in comparison to a reference population. Overall, MWS1.1 and MWS2.1 did not have an excess of genic copy number variation in their genomes (**Figure 1A**). Specifically, MWS1.1 had an ICNS score of 12 (**Supplementary Table S4**); MWS1.2 had a score of 3 (**Supplementary Table S5**); MWS1.3 had a score of 26 (**Supplementary Table S6**); MWS2.1 had a score of 7 (**Supplementary Table S7**); MWS2.2 had a score of 6 (**Supplementary Table S8**); MWS2.3 had a score of 73 (**Supplementary Table S9**). With a median ICNS of 9.5 for the reference cohort and a MAD of 7.413, MWS2.3 had a modified Z-score of 5.78, exceeding the 3.5 MAD threshold and therefore designating this individual as a major outlier (**Supplementary Table S3**). All other individuals’ modified Z-scores did not exceed the ± 3.5 outlier threshold **(Supplementary Table S3).** 58 of MWS2.3’s 73 ICNS points originated from CN events on chromosome 8. This is indicative of a major duplication event on that chromosome, as visualized in **Supplementary Figure S1.** However, none of the genes involved in this duplication event have been previously indicated to be relevant to the disease phenotype.

Furthermore, all individuals matched the reference CN averages for 21 of the 24 previously indicated genes relevant to the HSCR phenotype indicating no significant CN events in those regions, including *ZEB2*. The remaining 3 relevant genes (*KIF1BP, IKBKAP*, and *FAM213A*) did not have results, as they were not included in the 40,101 genes of the annotated CNPI ICNS reference location file. Both probands (MWS1.1 and MWS2.1) were not outliers for their ICNSs and do not have excess CN burden in comparison to the general population **(Figure 1A, Supplementary Figure S2 and S3).**

### RET Enhancer Risk Haplotype Transmission Implicates RET Dysregulation as a Modifier for HSCR in MWS1.1

We focused our analysis on a set of ten common noncoding enhancer variants at the *RET* locus because we have previously demonstrated that this specific variant set explains a large proportion of Hirschsprung disease risk through cumulative effects on *RET* regulatory output (Chatterjee et al. 2021). Building on this framework, we asked whether transmission of a high-risk RET enhancer haplotype could explain the presence of long segment HSCR in one MWS proband (MWS1.1), compared with a second proband with MWS but no HSCR (MWS2.1).

To address this, we analyzed the whole genome-sequencing data from the MWS1 and MWS2 trios and reconstructed parental haplotypes across the ten *RET* enhancer SNPs (rs788263, rs788261, rs788260, rs2506030, rs1547930, rs7069590, rs2435357, rs12247456, rs7393733, rs2505541) (**Supplementary Table S10**). Using trio genotypes, we inferred maternal and paternal transmission at each locus, phased the transmitted haplotypes, and assigned haplotype specific odds ratios (Chatterjee et al. 2021).

In the HSCR affected proband (MWS1.1) (**Figure 1A**), phasing revealed inheritance of a high-risk paternal *RET* enhancer haplotype (GCAGGTTGGT) with a haplotype OR of 11.01, whereas the maternally transmitted haplotype (CTGAATCACT) carried a substantially lower OR of 2.01. One variant, rs7069590, was missing from the trio VCF and was conservatively assumed to be homozygous reference in all three individuals for haplotype reconstruction. In contrast, in the proband with MWS but without HSCR (MWS2.1) (**Figure 1A**), transmission of the same haplotype from both parents (CTGAGTCGGC) was observed and that corresponded to a low-risk an OR of 1.33.

Given our prior functional studies showing that the *RET* enhancer risk haplotype produces a substantial reduction in RET regulatory output, these genetic observations are biologically consistent with a modifier effect in the MWS1.1 proband. In neural crest derived SK-N-SH cells, we previously demonstrated that this haplotype reduces *RET* expression by more than 50 percent through cumulative effects of the enhancer variants (Chatterjee et al. 2021). Complementary transcriptional analyses in human fetal gut further showed a similar reduction in RET expression in vivo associated with these noncoding haplotypes (Chatterjee et al. 2023). Together, these data support a model in which inheritance of the high-risk *RET* enhancer haplotype in MWS1.1 further lowers *RET* signaling in the context of ZEB2 haploinsufficiency, crossing a critical threshold required for enteric neural crest proliferation and migration and thereby contributing to the development of HSCR. These findings also underscore the importance of evaluating common enhancer variation at the level of haplotypes rather than individual SNPs, as variants with modest individual effects can collectively produce substantial reductions in gene expression and markedly increase disease risk.

### ZEB2 Variants and Pathogenicity

*ZEB2* encodes a transcription factor of the Zfh1 family of two-handed zinc finger and homeodomain proteins (Cacheux et al. 2001; Wakamatsu et al. 2001). It contains several well characterized functional domains, including a nucleosome remodelling and deacetylase interaction motif (NIM), a SMAD binding domain (SBD), a homeodomain (HD), a CtBP interacting domain (CID), and two zinc finger (ZF) clusters located in the N terminus (N ZF) and C terminus (C ZF) of the protein (Remacle et al. 1999). NIM and CID mediate recruitment of co repressors, whereas the ZF domains recognize CACCT(G) motifs in target promoters. Each ZEB2 ZF cluster comprises three C2H2 ZFs, and the N terminal cluster additionally contains a fourth CCHC ZF like motif (Remacle et al. 1999) (**Figure 1B**). Mapping of the variants detected in our probands showed us that the frameshift (p.(Lys284Asnfs*30)) lies on the N-terminal zinc-finger cluster whereas the (p.(Arg671*)) disrupts the Homedomain of ZEB2 (**Figure 1B**). Both variants are therefore predicted to disrupt critical DNA binding and transcriptional regulatory functions of the protein. To place these variants in the context of previously reported pathogenic variation, we queried *denovo-db*, a curated compendium of published human DNVs (Turner et al. 2017). Mapping of reported *ZEB2* variants onto the protein structure shows that DNVs associated with NDDs are broadly distributed across the coding sequence but are enriched in key functional domains, including the zinc finger clusters, SMAD binding domain, and homeodomain (**Figure 1B**). These variants include frameshift, missense, and stop gained mutations reported in individuals with developmental disorders, intellectual disability, autism spectrum disorder, and congenital anomalies. The two variants identified in our probands fall within regions of the protein that harbor multiple previously reported pathogenic DNVs, further supporting their functional relevance and consistency with established mutational patterns in ZEB2 related disease.

### Phenotypes of MWS1.1

Proband MWS1.1 presented with a complex phenotype involving multiple organ systems (**Figure 2**, **Supplemental Table S1**). Craniofacial features include an open mouth, pointed chin, uplifted earlobes, low-hanging columella, broad eyebrows, posteriorly rotated ears, prominent nasal tip, wide nasal bridge, and hypertelorism. Growth parameters indicate short stature and failure to thrive. Neurodevelopmental assessment revealed profound intellectual disability, poor speech, absent expressive language, delayed motor milestones including delayed ambulation, motor delay, and a broad-based gait. Additional neurological findings include impaired pain sensation, seizures, sleep disturbance, and suspected hypoplasia of the corpus callosum (reported but not confirmed by MRI). Behaviorally, the individual demonstrates a happy demeanor and stereotyped movements. Musculoskeletal examination identified genu valgum. Oromotor dysfunction was evident with drooling, and there was evidence of bowel and urinary incontinence. Genitourinary anomalies include cryptorchidism, chordee, bilateral vesicoureteral reflux (managed with ureteric reimplantation), and hydronephrosis, consistent with severe reflux nephropathy. Gastrointestinal involvement is notable for aganglionic megacolon (*long-segment Hirschsprung disease*) and chronic constipation. Cardiac evaluation revealed a dysplastic aortic valve with minor aortic regurgitation. Ocular findings include esotropia. Additionally, there was evidence of infantile and generalized hypotonia, as well as severe expressive and receptive language delay. In summary, the proband exhibits a multisystem disorder characterized by profound developmental impairment, distinctive craniofacial features, significant genitourinary and gastrointestinal abnormalities, and neurological and behavioural manifestations, including a happy effect and severe motor and language deficits.

**Figure 2:**
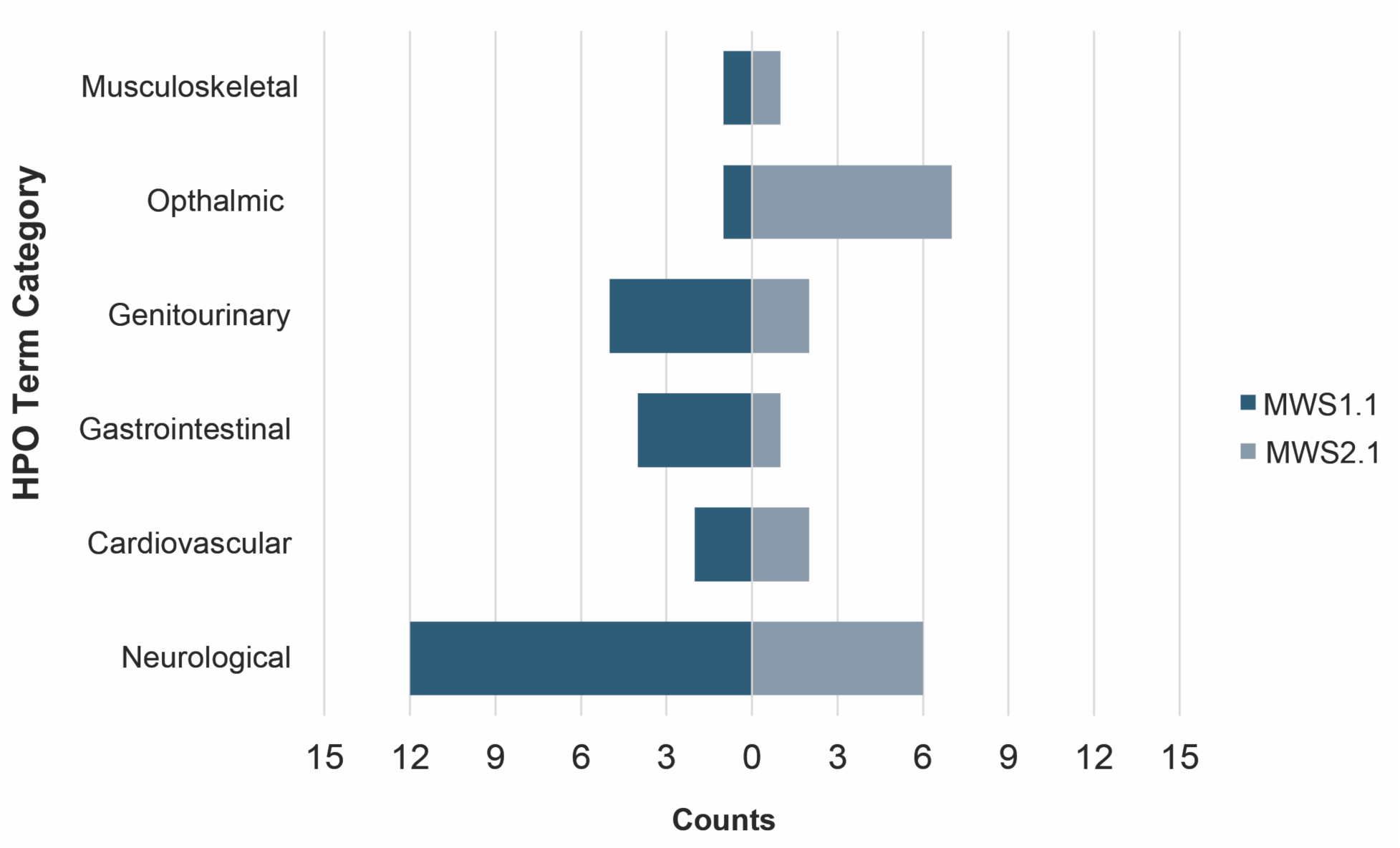
Human Phenotype Ontology (HPO)-based comparison of clinical features in the two Mowat Wilson syndrome probands. Counts of curated HPO terms for MWS1.1 and MWS2.1 grouped by major organ system categories, including neurological, cardiovascular, gastrointestinal, genitourinary, ophthalmic, and musculoskeletal phenotypes. Both individuals exhibit the characteristic multisystem involvement typical of Mowat Wilson syndrome, with neurological features representing the largest category in each proband. The distribution of HPO terms also highlights phenotypic differences between the two individuals, including additional gastrointestinal involvement in MWS1.1 consistent with the presence of Hirschsprung disease.

### Phenotypes of MWS2.1

Proband MWS2.1 exhibits a multisystem phenotype involving craniofacial, neurodevelopmental, ocular, cardiac, and renal abnormalities (**Figure 2**, **Supplementary Table S11**). Craniofacial examination revealed an open mouth, pointed chin, deeply set eyes, prominent nasal tip, low-hanging columella, broad eyebrows, posteriorly rotated ears, hypertelorism, and microcephaly, consistent with a distinctive craniofacial appearance. Growth parameters demonstrated short stature. Neurodevelopmental evaluation indicated profound intellectual disability with absent speech, impaired pain sensation, and a generally happy demeanor. Motor development was delayed, and gait was broad-based. A history of seizures was reported in early childhood, which subsequently resolved. Ophthalmologic assessment identified bilateral Axenfeld anomaly, cataracts, abnormal pupil morphology, and ectopia pupillae, resulting in significant visual impairment. Cardiac findings included a ventricular septal defect that did not require surgical intervention. Renal and genitourinary evaluation showed a single kidney, consistent with renal dysgenesis. Musculoskeletal examination was notable for long fingers (arachnodactyly) but no evidence of contractures, scoliosis, or other major skeletal deformities. Gastrointestinal history included constipation during childhood, which later resolved. In summary, the proband presents with a complex phenotype characterized by microcephaly, distinctive craniofacial features, bilateral ocular anomalies (Axenfeld anomaly and cataracts), profound neurodevelopmental impairment, single kidney, and a ventricular septal defect, accompanied by a happy effect and severe global developmental delay.

### ZEB2 and RET expression in human fetal enteric nervous system and developing neocortex

To place our WGS findings in a cellular context, we reanalyzed single cell RNA sequencing data from the developing human fetal gut (6–11 weeks post conception) generated by the Human Gut Cell Atlas (Elmentaite et al. 2021) (**Figure 3A**). In this dataset, ZEB2 transcripts are readily detected in enteric neural crest cell (ENCC) and glial progenitors, cycling ENCCs, and neuroblasts, with sustained expression across multiple differentiating neuronal branches, including inhibitory motor neurons, excitatory motor neurons, and intrinsic primary afferent/interneurons, and with appreciable levels in differentiating and mature glial clusters (**Figure 3B**). By contrast, RET expression is largely confined to neuroblasts and early neuronal branches, with highest expression in cycling neuroblasts and Branch B excitatory motor neuron populations and minimal expression in glial states (**Figure 3B**). These data, obtained from unaffected embryos, show that *ZEB2* and *RET* are co expressed in the same early ENS precursor and neuronal lineages at the developmental stages when neurogenesis is specified, providing a biological substrate for interaction between *ZEB2* haploinsufficiency and regulatory variation at *RET*.

**Figure 3.**
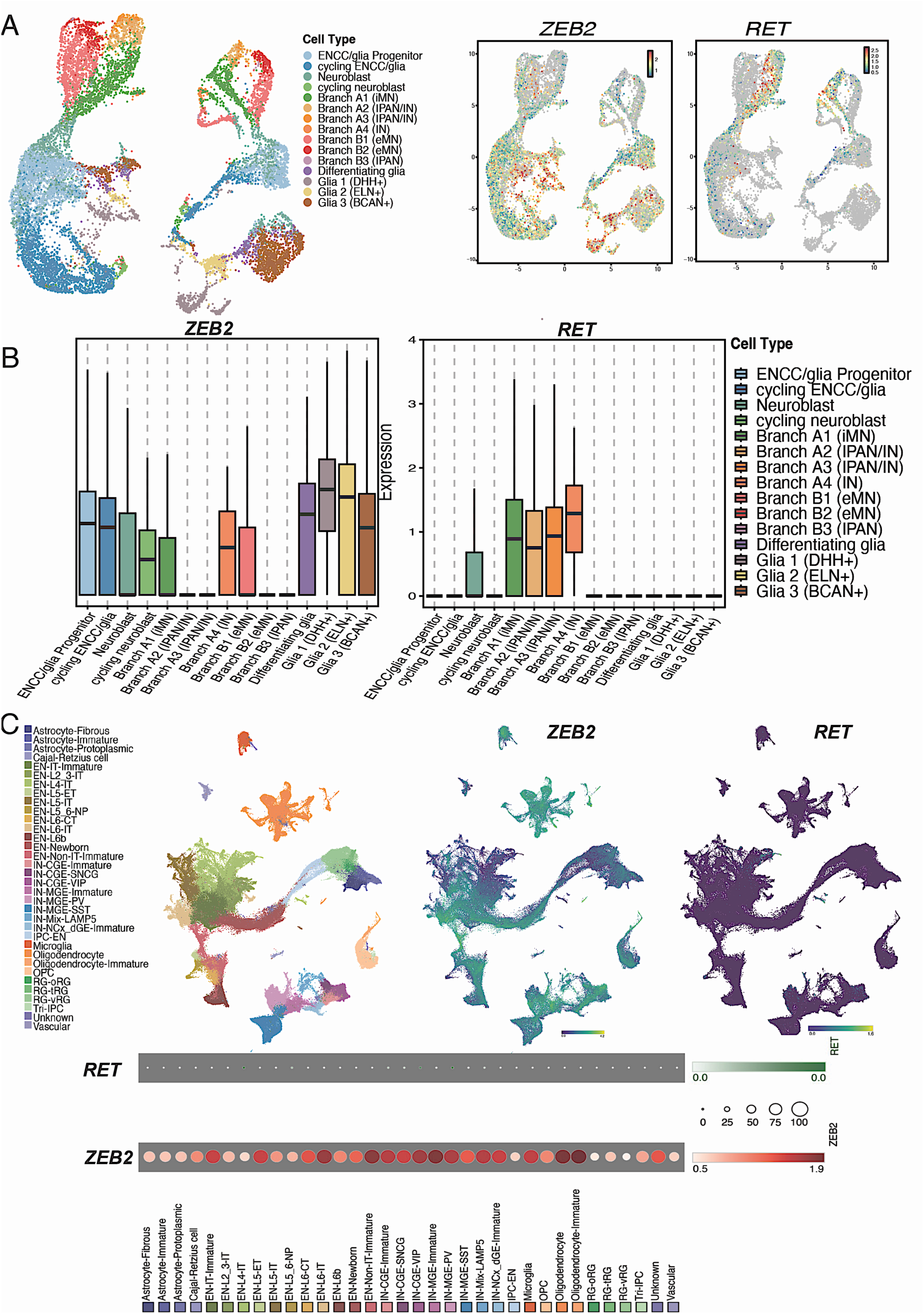
*ZEB2* and *RET* expression in the developing human enteric nervous system and neocortex. **(A)** UMAP representation of single-cell RNA sequencing data from the developing human fetal gut (6-11 weeks post conception) from the Human Gut Cell Atlas, showing annotated enteric neural crest-derived cell states including ENCC/glial progenitors, cycling progenitors, neuroblasts, multiple neuronal branches, and glial populations. Feature plots show expression of *ZEB2* and *RET* across these cell types. *ZEB2* is broadly expressed across progenitor, neuronal, and glial lineages, whereas RET expression is largely restricted to neuroblast and early neuronal populations. **(B)** Boxplots summarizing normalized expression levels of *ZEB2* and *RET* across ENS cell states. *ZEB2* shows sustained expression across multiple precursor and differentiated neuronal populations, as well as glial lineages, whereas *RET* expression is enriched primarily in neuroblasts and early neuronal branches with minimal expression in glial states. **(C)** UMAP visualization of single-cell RNA sequencing data from the developing human neocortex, showing annotated cortical progenitor and neuronal populations. Feature plots illustrate broad *ZEB2* expression across multiple cortical cell types, whereas *RET* expression is largely absent in the developing neocortex, indicating minimal overlap between the two genes in brain developmental lineages. These results highlight tissue-specific co-expression of *ZEB2* and *RET* in ENS progenitor populations but not in cortical lineages.

Because Mowat Wilson syndrome is characterized by prominent abnormalities of brain development, particularly involving the neocortex (Garavelli et al. 2017; Ivanovski et al. 2018), we next examined *ZEB2* and *RET* expression in the developing human fetal brain using single cell RNA sequencing data from a recently generated atlas of cortical development spanning early to mid-gestation, which profiles transcriptional states across diverse cortical progenitor and neuronal populations (Wang et al. 2025a). In contrast to the ENS, *ZEB2* is broadly expressed across multiple cortical progenitor and neuronal populations, whereas *RET* expression is largely absent in the developing neocortex, with minimal overlap between the two genes (**Figure 3C**). This tissue specific expression pattern suggests that reduced *RET* dosage, whether mediated by coding mutations or noncoding regulatory variants, is most likely to have functional consequences in the ENS, where RET operates within the same developmental lineages affected by *ZEB2* haploinsufficiency, rather than in the brain, where *RET* expression is largely absent.

Because *ZEB2* and *RET* are co expressed in early ENS precursor and neuroblast lineages, the data highlight a broader principle that may help explain variable penetrance of aganglionosis in MWS. Their shared expression domain suggests that MWS patients who develop Hirschsprung disease are likely to carry additional genetic variants in genes that operate within these neuronal precursor populations, the very cells that give rise to the enteric neurons affected in HSCR. This provides a rationale for future studies to prioritize modifier gene discovery within pathways and cell states active in ENS progenitors and neuroblasts, where combined effects of *ZEB2* haploinsufficiency and variants in other ENS developmental regulators may determine which individuals progress to aganglionosis.

## DISCUSSION

Mowat Wilson syndrome (MWS) is widely regarded as a prototypical monogenic disorder caused by heterozygous loss of function variants in *ZEB2*, yet marked variability in clinical presentation has been repeatedly documented (Garavelli and Mainardi 2007; Dagorno et al. 2020). Even among individuals with clearly pathogenic *ZEB2* variants, the presence and severity of gastrointestinal involvement, including HSCR, remains inconsistent. Large clinical series of molecularly confirmed MWS patients report HSCR in only a subset of affected individuals, typically ranging from 40 to 60 percent, underscoring that *ZEB2* haploinsufficiency alone does not fully determine enteric nervous system outcomes (Garavelli and Mainardi 2007; Dagorno et al. 2020). This variability has long suggested the existence of genetic or developmental modifiers, but systematic genome wide evaluation beyond the primary disease gene has been limited.

Multiple experimental and clinical observations support the plausibility that additional genetic modifiers influence the penetrance of HSCR in MWS. ZEB2, the gene mutated in MWS, encodes a transcription factor that plays a central role in neural crest development, including epithelial to mesenchymal transition and migration of neural crest cells that give rise to the enteric nervous system (Hegarty et al. 2015; Birkhoff et al. 2021). Experimental studies show that loss of ZEB2 disrupts neural crest delamination and migration, underscoring its essential role in early neural crest biology and peripheral nervous system formation (Charney et al. 2023).

Clinical observations further support this framework. HSCR associated with MWS is often indistinguishable from non-syndromic HSCR with respect to aganglionic segment length and histopathological features, suggesting convergence on shared developmental mechanisms rather than a syndrome specific enteric defect (Ishihara et al. 2005; Dagorno et al. 2020). Together, these findings support a model in which *ZEB2* haploinsufficiency creates a sensitized developmental background, while additional variants determine whether the threshold for enteric nervous system failure is crossed.

Within this framework, common regulatory variation at the *RET* locus represents a compelling genetic modifier influencing the manifestation of Hirschsprung disease (HSCR) in individuals with Mowat Wilson syndrome (MWS). *RET* dosage sensitivity is a well-established determinant of HSCR risk, with both rare coding mutations and common regulatory variants contributing to disease susceptibility (Emison et al. 2010; Chatterjee et al. 2016; Tilghman et al. 2019; Chatterjee et al. 2021). Early studies demonstrated that a common noncoding variant within a *RET* intronic enhancer increases HSCR risk by reducing *RET* transcription (Emison et al. 2005; Emison et al. 2010). More recent work has further demonstrated that several RET enhancers operate within a coordinated regulatory architecture, where combinations of variants collectively influence *RET* transcription during enteric nervous system development (Chatterjee et al. 2021). These variants cumulatively reduce *RET* expression during a critical developmental window required for enteric neural crest cell proliferation and migration. In individuals with *ZEB2* haploinsufficiency, the defining lesion in MWS, this reduction likely lowers the threshold for ENS developmental failure, providing a mechanistic explanation for why HSCR occurs in only a subset of MWS patients.

Earlier studies examined only the single *RET* enhancer SNP rs2435357 and reported no association with that SNP in MWS+HSCR patients (de Pontual et al. 2007). However, this approach did not capture the broader regulatory architecture of the locus, which was unknown at that time. Our analysis instead considers the full 10 SNP *RET* enhancer haplotype, consistent with current understanding that HSCR susceptibility arises from the combined effect of multiple regulatory variants (Chatterjee et al. 2016; Chatterjee et al. 2021). These findings underscore the importance of evaluating common enhancer variation at the level of haplotypes rather than individual SNPs, as variants with modest individual effects can collectively produce substantial reductions in gene expression and markedly increase disease risk.

These findings have broader implications for both genomic diagnostics and disease modelling. Clinically, they suggest that whole genome sequencing can provide additional interpretive value even after identification of a pathogenic Mendelian variant, particularly for syndromes with variable expressivity and multisystem involvement. Biologically, they highlight the importance of studying disease genes within their regulatory and developmental networks rather than in isolation. More broadly, these observations align with a growing body of work in neurodevelopmental disorders showing that conditions traditionally viewed as monogenic often reflect interactions between high impact coding variants and a background of common genetic risk (Niemi et al. 2018; Oetjens et al. 2019). This concept is consistent with a wider literature on Mendelian disease demonstrating that modifier loci and genetic background can substantially influence penetrance and phenotypic variability (Cooper et al. 2013).

Future studies in larger, deeply phenotyped MWS cohorts will be required to more precisely quantify the penetrance modifying effects of *RET* and other neural crest development pathways, and to incorporate additional classes of variation detectable by whole genome sequencing, including structural variants. Integrating these genomic data with emerging cell type resolved developmental atlases of the enteric nervous system will help prioritize modifiers that act within the same temporal and cellular contexts as *ZEB2*. More broadly, our findings support an integrative model in which Mendelian diagnoses serve as anchors within a wider genomic landscape, where both rare and common variants collectively influence developmental outcomes. This framework provides a path toward understanding the genetic basis of variable expressivity in syndromic neurocristopathies and highlights the importance of incorporating regulatory variation and modifier effects into both mechanistic studies and future genomic diagnostics.

## METHODS

### Mowat Wilson Syndrome Families

Two families were identified with Mowat Wilson Syndrome (MWS) including one with a child with MWS and Long-Segment HSCR (Family MWS1) and one with MWS but not HSCR (Family MWS2). DNA and phenotypes were collected at the Sydney Children’s Hospitals Network and were approved for sequencing by Human Research Ethics Committee (SCHN HREC) and Sydney Children’s Hospitals Network Research Governance Office (SCHN RGO) under approval numbers 2019/ETH12990; 2020/STE05398 respectively. Details of their phenotypes are in **Supplementary Table S11**.

### Whole-Genome Sequencing

Two families with Mowat Wilson Syndrome were sequenced at the New York Genome Center on an Illumina sequencer to a coverage of 30x (**Supplementary Table S12; Supplementary Figure 4**). Post-sequencing, reads were aligned to the GRCh38 version of the human genome using BWA-MEM (Li et al. 2009). Copy number variation across the genome was assessed with QuicK-mer2 (Shen and Kidd 2020) and CNPI (Ustanik and Turner 2025), while insert size and coverage metrics were computed using Picard (https://broadinstitute.github.io/picard/). Structural variants were identified with both *Manta* (Chen et al. 2016) and *Delly* (Rausch et al. 2012). Single-nucleotide variants (SNVs) and insertions/deletions (indels) were called on a per-individual basis using *DeepVariant* (Poplin et al. 2018), and jointly genotyped across the trio using *GLnexus* (Yun et al. 2021). Finally, DNVs were detected with *HAT* (Ng and Turner 2024). Altogether, comprehensive variant discovery was pursued across the dataset. Variants were prioritized based on HPO terms using *Exomiser* (Smedley et al. 2015). Variants were also compared to known DNVs from the literature (Turner et al. 2017; Wang et al. 2025b) (**Supplementary Table S13**) and plotted using *plot-protein* (Turner 2013).

### Copy Number Burden

To determine if any individuals had genic deletions or duplications, we utilized CNPI (described above). CNPI was run on an annotated reference location file consisting of 40,101 genes, obtained from the publicly available CNPI GitHub (https://github.com/TNTurnerLab/CNPI). CNPI was run with conservative, chromosome-wide CN thresholds of 1.3 for deletions and 2.7 for duplications, as described previously (Ustanik and Turner 2025). The outputs from this analysis included chromosomal sex determination, digital karyotyping, genotyping, and statistical analysis of CN values for each of the 40,101 genes in every individual.

Next, we utilized the Individual-Level Copy Number Score (ICNS) feature of the CNPI toolkit to quantify the CN burden in each individual using their CNPI-generated genotype results. Per-gene scores were determined based on the upper (99.9^th^) and lower (0.01^th^) percentiles of CNPI-derived CN averages obtained from the 1000 Genomes Project, also publicly available on the CNPI GitHub. Cumulative ICNSs were also generated for each sample. Again, conservative thresholds of 1.3 and 2.7 for deletions and duplications, respectively, were used for this analysis. To account for the impact of sex chromosomes on individuals’ scores, we utilized the appropriate adjustment features provided by the ICNS program as well as the male and female reference percentile files available on the CNPI GitHub.

To identify potential outlier ICNS values, modified Z-scores *(M_i_)* were computed for each individual using the sample median (*x̃*) and the median absolute deviation *(MAD).* This robust approach was used because the sample was small and non-normally distributed. Observations with | *M_i_* | > 3.5 were classified as potential outliers, following (Iglewicz and Hoaglin 1993).

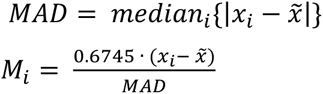

To compare the ICNSs, from each individual, against those of an unaffected population, data derived from Supplementary Table 16 of Ustanik and Turner (2025) were used to generate an ICNS density plot in R using the ggplot2 package (https://cran.r-project.org/web/packages/ggplot2). Finally, to visualize CN events on a chromosomal level, the CNPI plotting function was utilized with the same respective 1.3 and 2.7 deletion and duplication CN thresholds. For MWS2.3 specifically, chromosome 8 was visualized from a start position of 63712 and a stop position of 145006046, to visualize the duplication present in their CNPI-generated genotype output.

### RET Haplotype Prediction

We assessed the haplotype risk of 10 enhancer variants across *RET* (Chatterjee et al. 2021) as follows from joint-genotyped family variant call files. First, we assess the genotypes of the ten specific *RET* SNPs on hg38. If a site is missing genotypes, we assume each person is homozygous for the reference allele. Second, for each SNP, we use Mendelian logic to figure out which allele in the child came from the mother and which came from the father. Third, we label each transmitted allele as risk (R) or non-risk (N) based on hard-coded risk bases (Chatterjee et al. 2016; Chatterjee et al. 2021; Chatterjee et al. 2023). Finally, in a fixed SNP order, based distance from *RET* transcription start site, we build the maternal and paternal haplotype sequences and then assign the odds ratio (OR) for each haplotype from the *RET* locus we have previously identified from genetic and functional studies of HSCR (Chatterjee et al. 2021; Chatterjee et al. 2023).

### Single Cell RNA-seq Data

The processed AnnData file for 62,849 cells isolated from 9 donor GI tract from 6-11 weeks post-conception developing human gut was downloaded from https://www.gutcellatlas.org/ (Elmentaite et al. 2021). This was converted to a Seurat object using the “Convert” function in *Seurat* (Stuart et al. 2019). We utilized SCT transform in Seurat to normalize the samples to retain cells with counts >100 UMI and detected in >10 cells in all samples. We used the authors’ described 10 principal components to generates the Uniform Manifold Approximation and Projection (UMAP) embedding on these cells using the python package “umap-learn” in *Seurat*. The cell identity of the UMAP clusters was derived from the metadata file provided by the authors. Human fetal brain single cell RNA-seq was downloaded from https://cell.ucsf.edu/snMultiome/build1/ with author embedded annotations and cell labels. Details of the study can be found here- (Wang et al. 2025a)

## Data Availability

Data supporting this study are available from the corresponding author upon reasonable request.

## Ethics approval and consent to participate

Phenotypes were collected at the Sydney Children’s Hospitals Network and were approved for sequencing by Human Research Ethics Committee (SCHN HREC) and Sydney Children’s Hospitals Network Research Governance Office (SCHN RGO) under approval numbers 2019/ETH12990; 2020/STE05398 respectively

## Availability of data and materials

Data supporting this study are available from the corresponding author upon reasonable request.

## Author Contributions

SCh and TNT conceived and designed the study. RP and DM consented and collected patient phenotypes and DNA. SC, IB, TNT and SCh analysed the data. SC, TNT and SCh wrote the manuscript. All authors approved the final version of the manuscript.

## Declaration of Interests

The authors declare no competing interests.

## Funding

SCh is supported by a NIDDK R01 award DK135089, NICHD R03 award HD116004 and The Maci Whisner Research Grant from Mowat Wilson Disease Foundation. TNT is supported by NIMH R01 award MH126933 and NICHD R03 award HD116062. The funders had no role in design of the study or data interpretation.

## Acknowledgements

We sincerely thank the individuals with Mowat Wilson syndrome and their families for their generous participation and support of this study. We are also deeply grateful to the Mowat Wilson Syndrome Foundation for their continued support and assistance, which made this work possible.

## SUPPLEMENTARY FIGURES

**Supplementary Figure 1.**
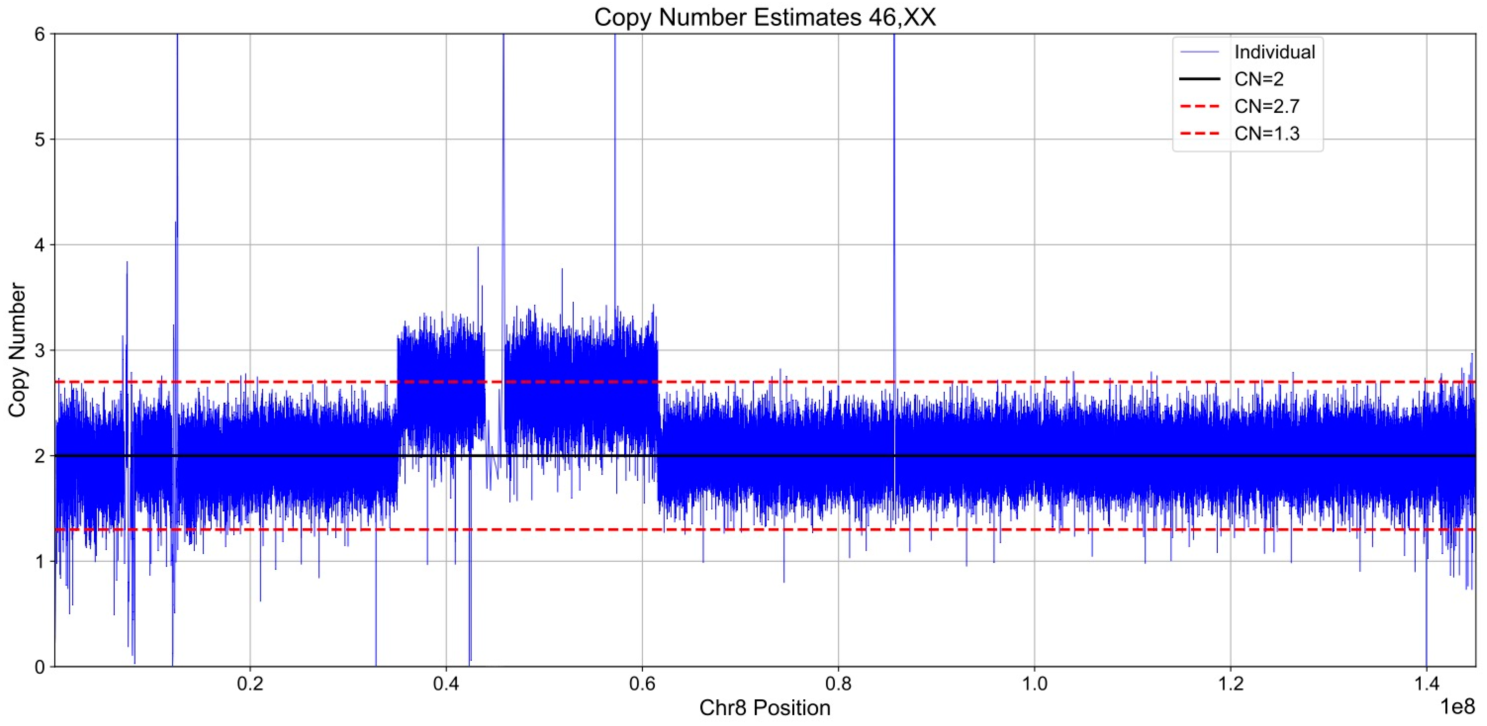
MWS2.3 chromosome 8 copy number plot.

**Supplementary Figure 2.**
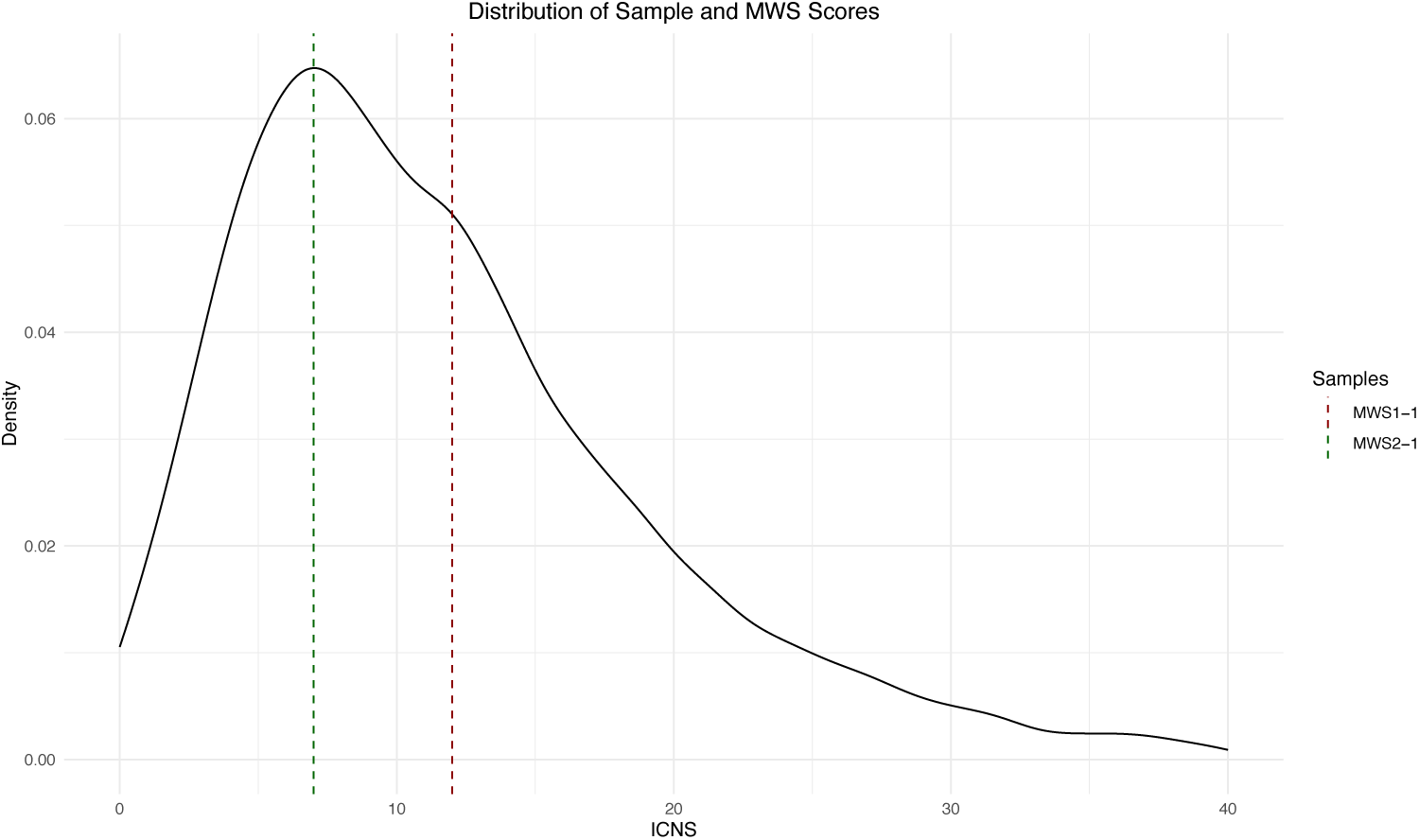
ICNS density plot of MWS1.1 and MWS2.1.

**Supplementary Figure 3.**
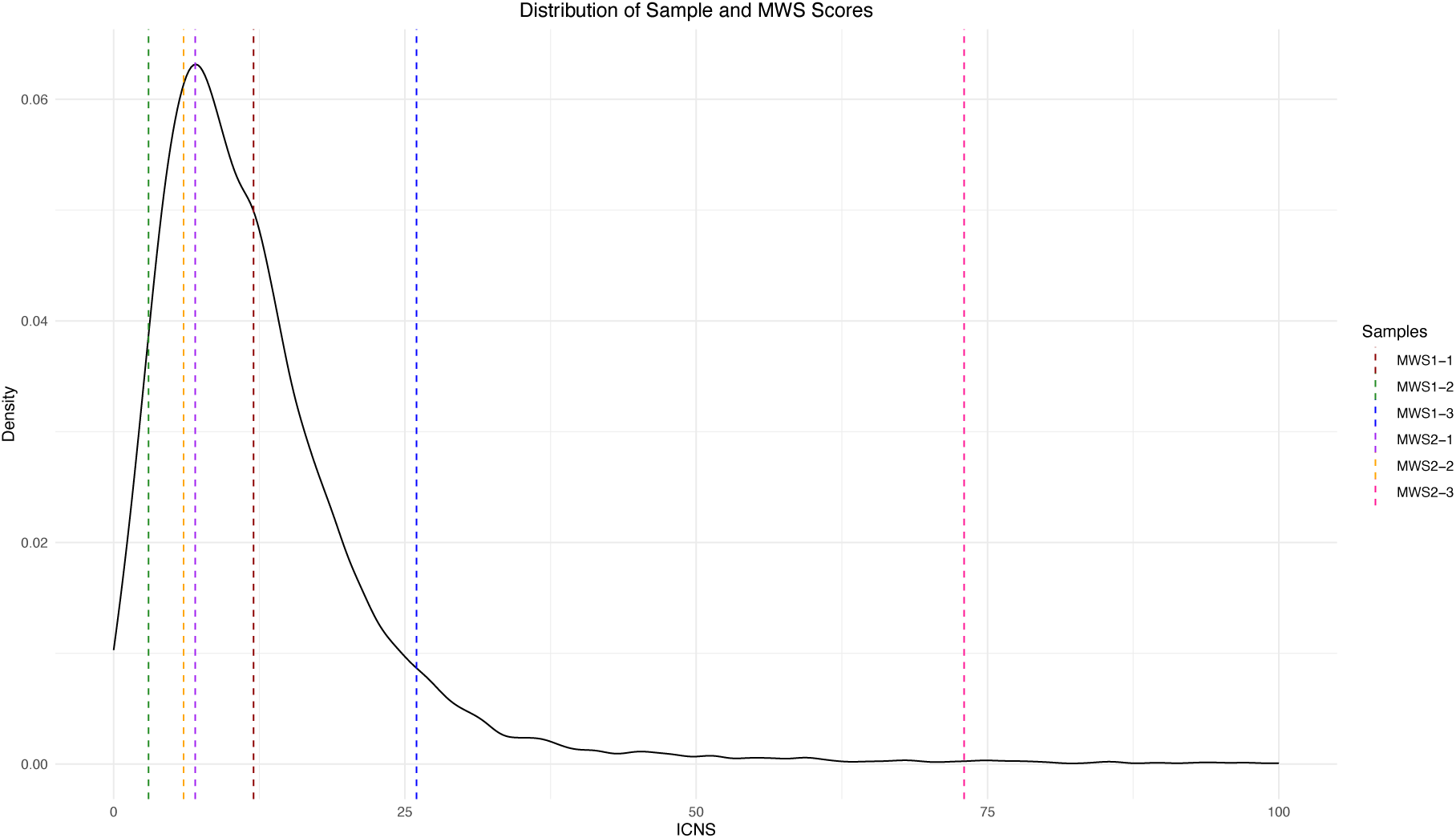
ICNS density plot of the 1000 Genomes Project and all six family members.

**Supplementary Figure 4.**
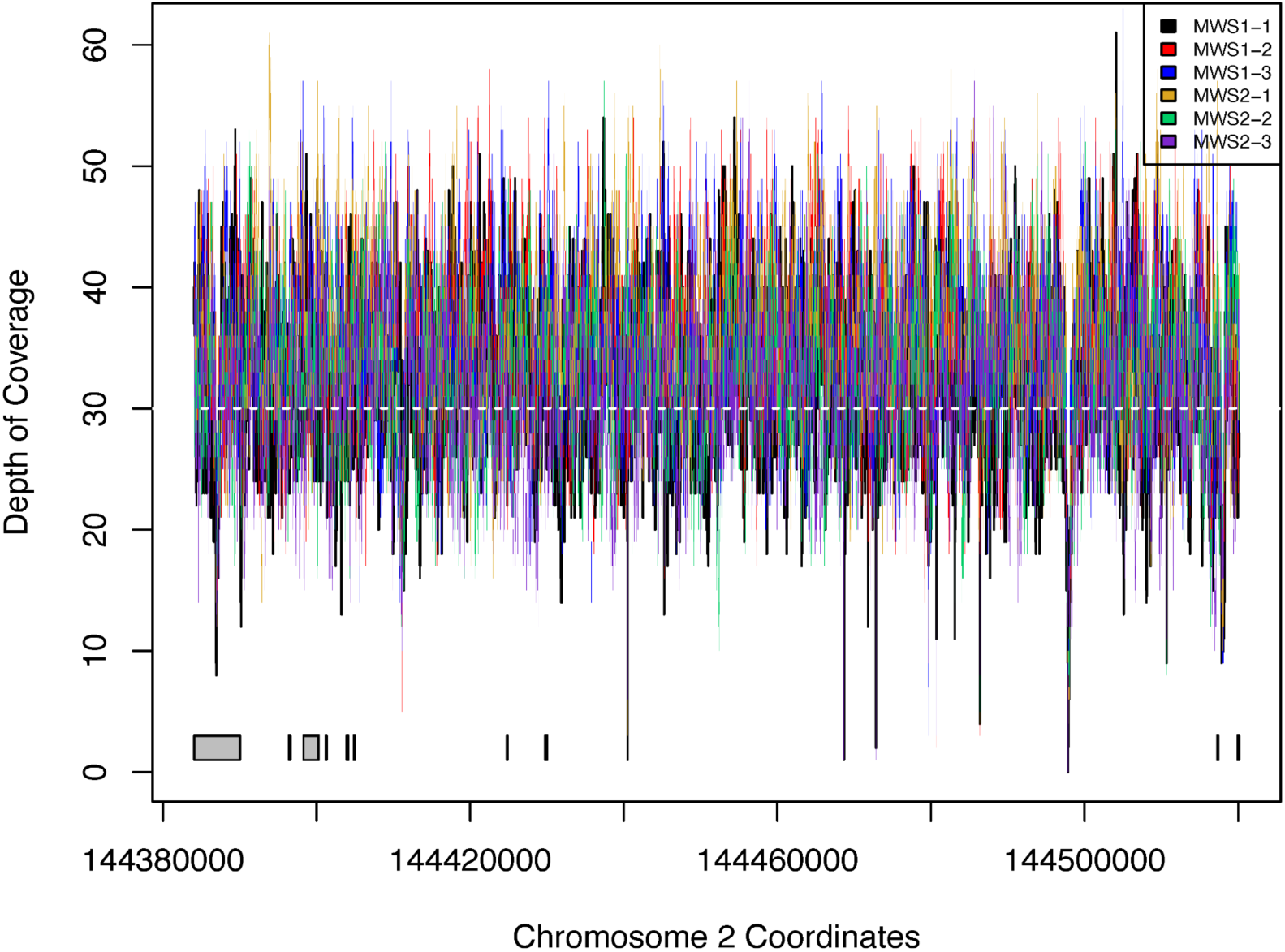
WGS coverage over *ZEB2* (exons are grey rectangles at the bottom)

## Notes

### Competing Interest Statement

The authors have declared no competing interest.

### Author Declarations

DNA and phenotype were collected at the Sydney Childrens Hospitals Network and were approved for sequencing by Human Research Ethics Committee (SCHN HREC) and Sydney Childrens Hospitals Network Research Governance Office (SCHN RGO) under approval numbers 2019/ETH12990 and 2020/STE05398 respectively

